# Failure of the New Kidney Allocation System in a Specific Ethnic Group

**DOI:** 10.1101/2020.11.26.20239236

**Authors:** Texell Longoria-Dubocq, Yaritza Pizarro-Gonzalez, Isabel Mayorga-Perez, Mariel Javier-Gonzalez, Pedro Hernandez-Rivera

**Author notes:** **Contact Information (Texell Longoria):** Address: Department of Surgery, University of Puerto Rico, Medical Sciences Campus, PO BOX 365067, San Juan, PR 00936-5067. **Disclosures:** The authors declare that they do not have any relevant or material financial interests that relate to the research described in this paper.

## Abstract

**Introduction:** The Kidney Allocation System (KAS) implemented on December 4, 2014, was expected to improve kidney transplant list wait-time and allocate more kidney to high cPRA patients. This study aims to demonstrate outcomes after the implementation of the KAS in a Hispanic transplant center.

**Methods:** Retrospective study from a prospectively maintained database from a single-transplant center. Included all DDKT from July 2013 to June 2016. Compare and analyze DDKT selection and outcomes before and after KAS implementation.

**Results:** The overall number of kidney transplants performed during this period was 220. All of the patients were Hispanic. Included 50.5% Pre-KAS and 49.5% Post-KAS. Pre-KAS group had a significantly shorter waiting-time list than the post-KAS group, 900.05 vs 1126.75 respectively. EPTS less than 20% significantly improved in the Post-KAS group compared to the Pre-KAS group, 41.3% vs 20.9% respectively. No differences observed in KDPI, 1-year graft failure, or 1-year mortality rates.

**Conclusion:** This might be the first Hispanic only cohort evaluating the effects of KAS on a moderate volume kidney transplant center. The new system increased the wait-time list by approximately 25%, and it did not improve graft quality, graft failure, or mortality rates.

## 1. Introduction

Kidney transplantation is the gold standard and most cost-effective treatment for patients with end-stage renal disease (ESRD). Especially, if it is performed before commencing long-term dialysis. Each additional year in dialysis treatment worsens patient survival. However, due to a shortage of available organs and long waiting list times, only 2.5% of patients with ESRD undergo transplantation as initial renal replacement therapy.^1^

The original kidney allocation program was implemented in 1987 with the formation of the United Network for Organ Sharing (UNOS). It was based on the HLA system, time on the waiting list, and geographic location. Increased demand for kidney transplants along with the decreased number of kidney donors resulted in a longer wait-list time and a pronounced disparity in wait-list time between different geographic locations and ethnicities. Donor’s kidneys were classified as standard criteria donor (SCD) or expanded criteria donor (ECD). When compared to SCD donors, ECD kidneys had a 70% increased risk for graft failure. Nevertheless, patients that received ECD grafts had better outcomes than matched dialysis-treated patients.^2^ However, this system did have several flaws. Older patients would receive a kidney and probably die with a functioning graft, while younger patients with a higher survival probability would receive a graft with poorer quality leading to a second transplant. Also, minority groups and highly sensitized patients would have longer wait-list time compared to their white and unsensitized counterparts.^3^ These issues with the old system demanded improvement in the allocation of kidneys by using more objective measures to avoid these disparities.

In 2005, UNOS authorized the revision of the KAS. Their goal was to increase access to transplants for candidates who were harder to match due to high Calculated Panel-Reactive Antibody (cPRA), minimize inequalities due to how the waiting time was calculated, and minimize unrealized life years post-transplantation, re-transplant rates and the rate of discarded kidneys. The new KAS, implemented on December 4, 2014, began using the Kidney Donor Profile Index (KDPI) and Expected Post Transplant Survival (EPTS). The smaller the number of KDPI and EPTS, the better graft quality and recipient expected survival, respectively. Under the revision of the KAS, highly sensitized candidates (those with a cPRA of 80% or greater), also have national and regional priority, receiving additional points which reflect their biological degree of sensitization.^4^

Recently, the average wait-time, once a person is added to the national organ transplant waiting list, is from three to five years.^5^

According to a short-term study from Sanchez et al, the implementation of the new KAS has resulted in a more equitable distribution of kidney transplants; there was a decrease in transplants for Caucasians and an increase for Hispanics and African Americans.^6^ Other studies suggest the new system has resulted in a higher proportion of transplants in patients with high cPRA with higher quality grafts.^7^

Implementation of the new KAS was aimed to decrease waitlist time and increase the number of transplants in high volume centers.^8^ Research evaluating such effects on low to moderate volume transplant centers have been scarcely reported.^9^ Our aim is to demonstrate how the implementation of the KAS influenced kidney transplant outcomes in a predominantly Hispanic population.

## 2. Methods

### 2.1 Selection process

A retrospective study evaluating all medical records of kidney transplants performed in the only transplant center in Puerto Rico, before and after the implementation of KAS. The Auxilio Mutuo Hospital is a multiorgan transplant center located in Region 3 of UNOS. The program is composed of four kidney transplant surgeons. Inclusion criteria were met if the patient underwent a deceased-donor kidney transplant (DDKT) between July 2013 and June 2016 with a post-operative follow up of 1-year. Living-donor kidney and multiorgan transplants were excluded since KAS criteria are only applicable to DDKT. To compare effects of KAS on DDKT volume and outcomes, patients were divided into pre-KAS (if transplant occurred before December 4, 2014) and post-KAS (if transplants occurred during or after December 4, 2014) which account for a total of 3 years (1.5 years pre-KAS and 1.5 years post-KAS).

### 2.2 Kidney Donor Profile Index (KDPI)

All of the deceased donor grafts were entered into the Organ Procurement and Transplant Network (OPTN) calculator in order to determine each graft’s KDPI using 2017 as a control. KAS added the KDPI score to the kidney allocation formula after December 4, 2014. All cases prior to KAS did not have a KDPI score to compare graft quality against the new cases. KDPI median pre-KAS and post-KAS was calculated using descriptive statistics. Then, patients were divided into three groups depending on their KDPI score: group 1= 0-20%, group 2= 21-85%, and group 3= 86-100%. A lower KDPI score is associated with longer graft survival.^10^ According to the OPTN KDPI guide, each group has a 1 year estimated graft survival as follows: Group 1 96.5%, Group 2 89.6% to 96.4%, and Group 3 81.8% to 89.5%.^4^

### 2.3 Estimated Post-Transplant Survival (EPTS)

The patient’s past medical history and wait-time list or dialysis start date were entered into the OPTN EPTS calculator in order to determine the EPTS score for DDKT patients that took place pre-KAS. EPTS was also added to the kidney allocation formula after KAS implementation. Median EPTS was calculated using descriptive statistics. Also, EPTS was categorized into EPTS 0-20% and EPTS 21-100%. A lower EPTS score is associated with better 10-year survival.^4^

### 2.4 Wait-time list

We recorded time on the kidney transplant list using the date in which the patient was placed on the list or dialysis start date, whichever came first. When the patient was successfully transplanted he or she was removed from the list. If a patient had a prior kidney transplant prior to the observed period, the wait time list started when the patient was placed in dialysis.

### 2.5 Calculated Panel Reactive Antibodies (cPRA)

Recipient cPRA was obtained from medical records. A cPRA equal or above 80 would improve the recipient’s chances of being transplanted, according to the OPTN algorithm implemented by the KAS.^11^ Therefore, highly sensitized patients were defined as patients having a cPRA score equal to or above 80.

### 2.6 Statistical analysis

Statistical analyses were performed using SPSS Software version ##(IBM, Chicago, IL). Categorical values such as gender, KDPI group, EPTS group, and cPRA group were compared and analyzed by Pearson’s Chi-Square. Comparisons for continuous variables such as age, BMI, and wait-list time were analyzed by Levene’s Test. A p-value of <0.05 was considered statistically significant.

## 3. Results

### 3.1 Demographics

A total of 219 patients were included in this study. All of them were of Hispanic ethnicity. Both Pre-KAS (50.68%) and Post-KAS (49.77%) groups had a similar number of kidney transplants during the studied period. The implementation of the KAS significantly decreased the patient’s median transplant age from 53 to 47.67 years old. However, there was no difference in gender, BMI, cPRA, or blood type distribution between KAS groups. (Table 1).

**Table 1.** Pre-KAS and Post- KAS transplant recipient demographics.

|  | *Pre-KAS (n=111)* |  | *Post-KAS (n=109)* |  | *p-value* |
| --- | --- | --- | --- | --- | --- |
| ***Age*** | 53 | ± 1.22 | 47.67 | ± 1.29 | **0.003** |
| ***Wait-time*** | 900.05 | ±65.84 | 1,126.75 | ±86.61 | **0.038** |
| *Male* | 71 | 64.0% | 70 | 64.2% | 0.968 |
| *Female* | 40 | 36.0% | 39 | 35.8% |  |
| ***BMI*** |  |  |  |  | 0.085 |
| *Underweight* | 2 | 1.8% | 0 | 0.0% |  |
| *Normal* | 31 | 27.9% | 18 | 16.5% |  |
| *Overweight* | 42 | 37.8% | 52 | 47.7% |  |
| *Obese* | 36 | 32.4% | 39 | 35.8% |  |
| ***Blood Type*** |  |  |  |  | 0.284 |
| *A* | 51 | 45.9% | 40 | 36.7% |  |
| *B* | 6 | 5.4% | 13 | 11.9% |  |
| *AB* | 3 | 2.7% | 4 | 3.7% |  |
| *O* | 51 | 45.9% | 51 | 46.8% |  |
| *cPRA (mean)* | 10.44 | ±1.57 | 9.48 | ±1.27 | 0.352 |
| *KDPI (mean)* | 39.86 | ±2.78 | 42.72 | ±2.66 | 0.459 |
| ***EPTS (mean)*** | 49.23 | ±2.8 | 40.27 | ±3.1 | **0.033** |

### 3.2 KAS Outcomes

The benefits of the new KAS could not be appreciated at this institution. Surprisingly, the Post-KAS group had a significantly longer wait-time list compared to Pre-KAS, 1126.76 (±86.61) vs 900.05 (±65.84) respectively (Table 1). The mean EPTS score significantly differed between the Pre-KAS and Post-KAS groups, 49.23 (±2.8) vs 40.27 (±3.1) (Table 1). Post-KAS groups had a significantly higher number of EPTS less than 20% compared to their Pre-KAS counterparts, 41.3% vs 20.9% respectively (Figure 1). However, there was no difference between both groups in relation to KDPI mean or categories (Figure 2). Graft failure and mortality rates at 1-year post-transplant were not affected by the new system (Figure 3).

**Figure 1.**
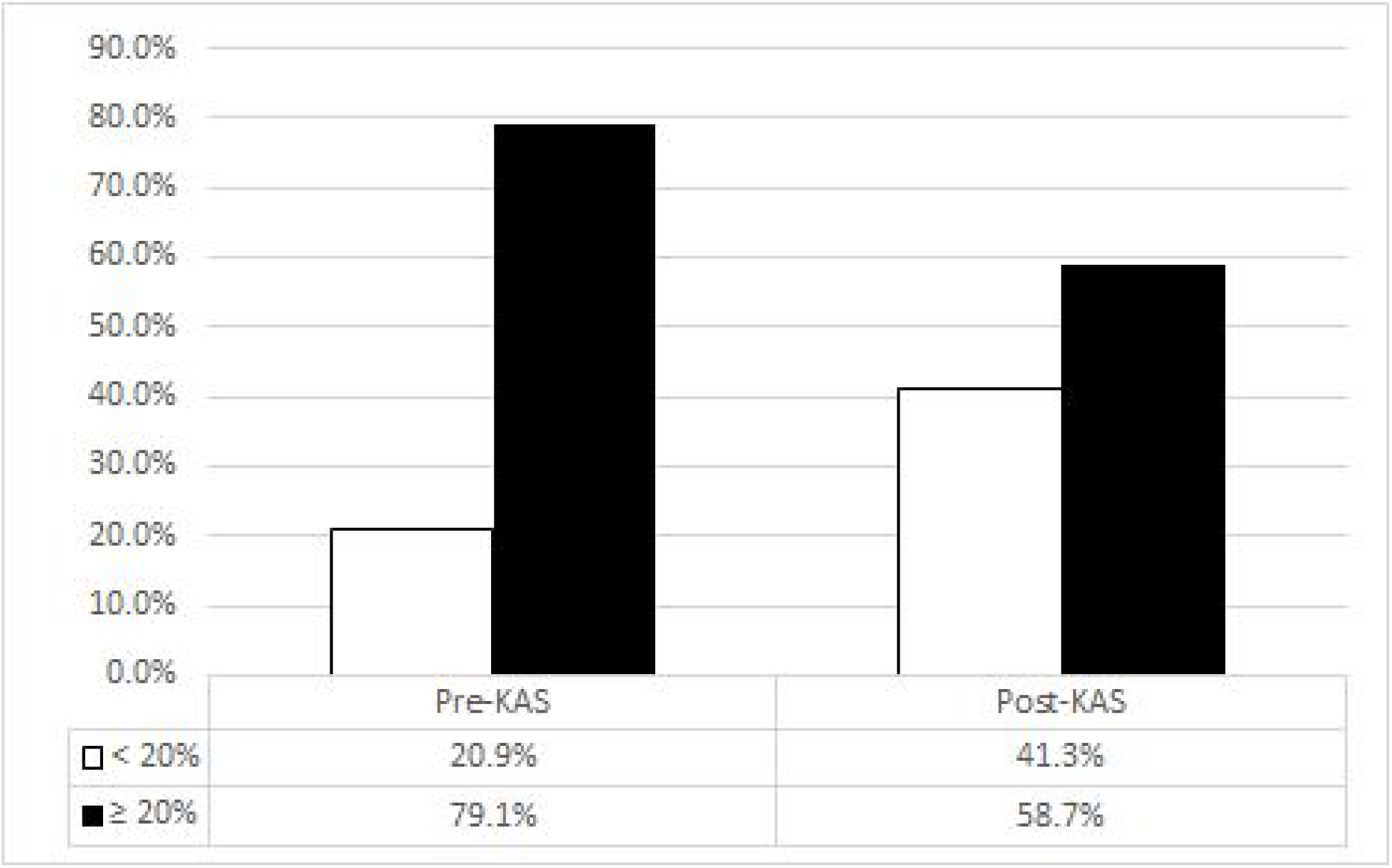
KAS influence on Estimated Post-Transplant Survival (EPTS) distribution. *p-value <0*.*001*.

**Figure 2.**
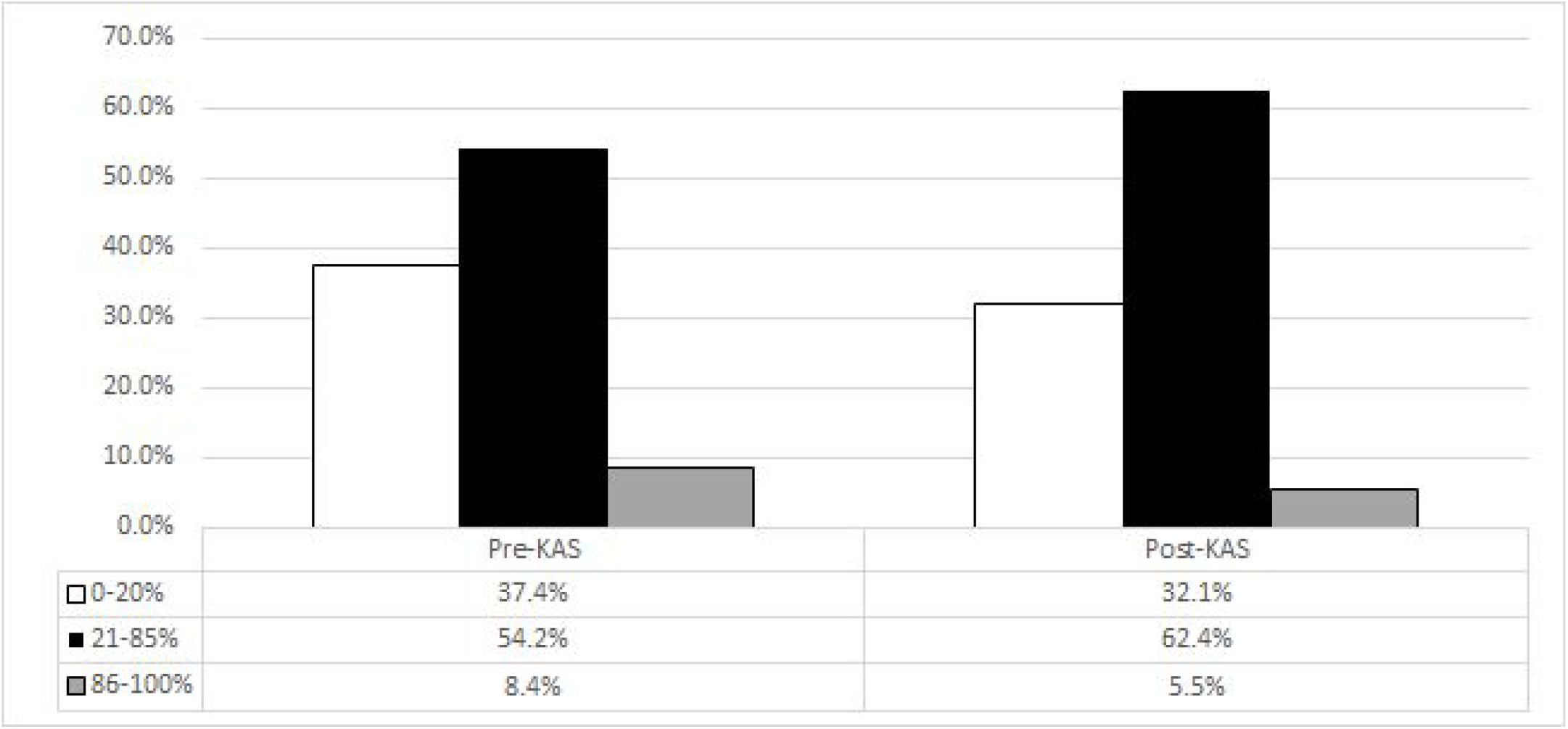
KAS influence on graft quality distribution and Kidney Donor-Profile Index (KDPI) before and after implementation. *p-value 0*.*426*

**Figure 3.**
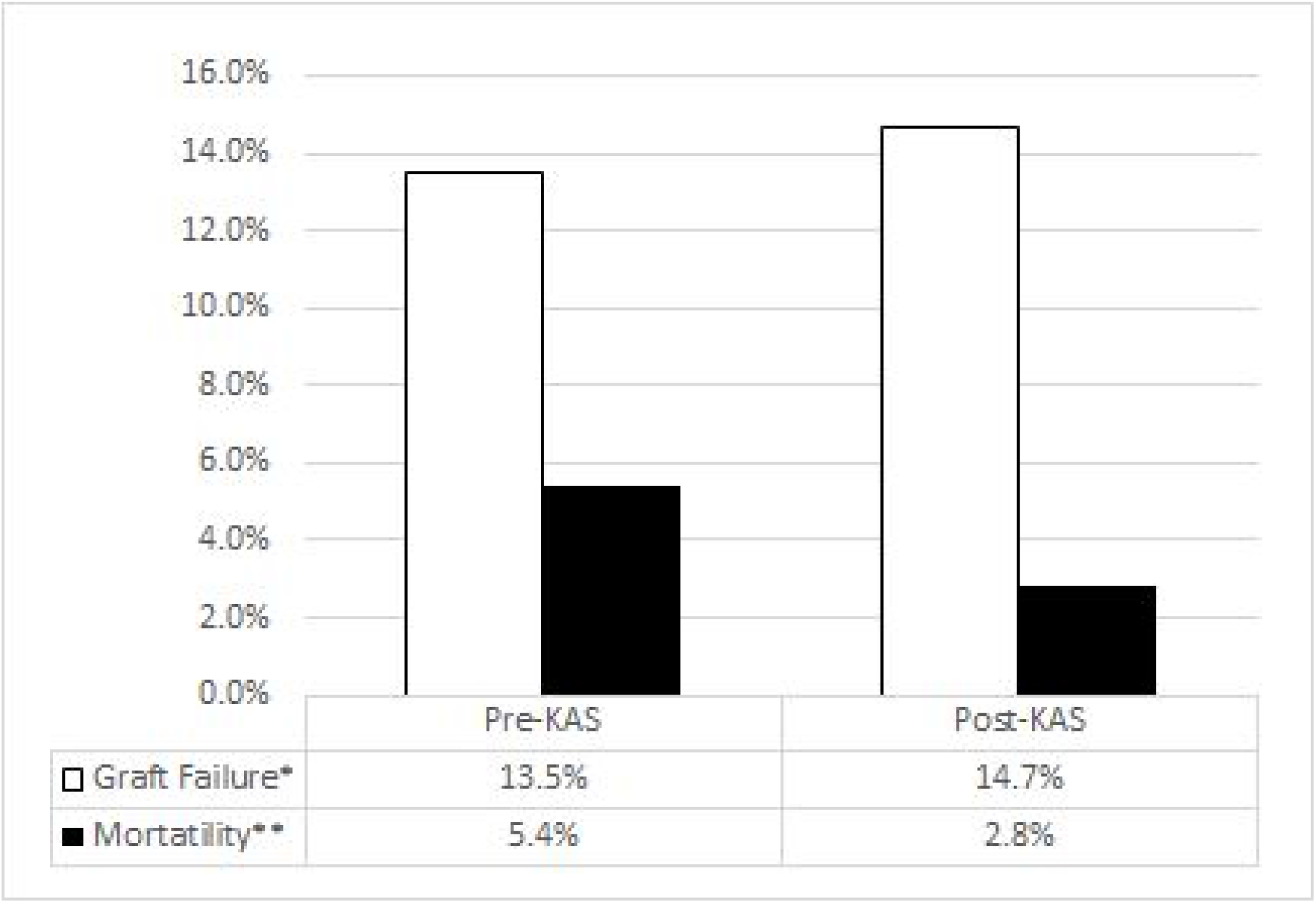
Recipients graft failure and mortality at 1-year post-transplant. *p-value=0*.*804*\**; p-value=0*.*321*\*\*

## 4. Discussion

Auxilio Mutuo Hospital is classified as a medium-volume transplant center.^9^ Our findings demonstrate that KAS did not benefit medium volume institutions with a Hispanic population. Although, there was a better selection of patients by using the EPTS score after KAS implementation. The quality of the donor graft allocation, wait-time list, graft failure rate, or mortality rate did not improve in the 1-year post-transplant period.

Transplantation volume rates were not affected by KAS. However, studies from high volume centers demonstrated that within minorities, specifically African Americans and Hispanics, rates of transplantation had increased after KAS.^6^, ^12^ Our institution did have the same results. Even though our kidney transplant list is composed of mostly, if not all, Hispanics. Locally there are no other transplant centers, but we do compete with other centers from Region 3. The closes one being over 1,000 miles away.

Another issue that remains to be study is how this new system has affected low to medium volume transplant centers, in terms of receiving organs. Studies from high volume centers have demonstrated an increase in kidney transplants and decreased wait-time list after the implementation of the KAS.^13^ At our center, similar changes were not observed. An even though recipients were more objectively selected with the EPTS, at a 1-year post-transplant period it did not change outcomes.

Therefore, further studies need to evaluate the effects of KAS on the different types of transplant centers taking into account volume and wait-time list census. There were several limitations encountered with this retrospective approach. The period studied was relatively short due to follow-up constraints, and matching time periods. Also, changes in types of immunosuppressant medication in earlier years could not be analyzed or compared due to a lack of information.

This might be the first completely Hispanic cohort study, more specifically Puerto Ricans. This means that other studies include Hispanics as a whole without emphasizing their heritage, which has been demonstrated to social and health disparities.^14^ Future studies evaluating the Hispanic heritage could shed some light on social and health disparities within this large ethnic group.

### 4.1 Conclusion

The new system did not improve transplant volume of highly sensitized patients, wait-list time, graft quality, graft failure rates, or mortality rates. Although EPTS was improved compared to the previous system, as a whole, the implementation of the KAS did not reach its goals at this moderate volume Hispanic only institution. Wait-time list increased by 25%, transplant volume remained stagnant, as well as graft quality and failure. This is the only cohort evaluating the effects of KAS on DDKT outcomes in a completely Puerto Rican population.

## Data Availability

Raw data were generated at Auxilio Mutuo Hospital. Derived data supporting the findings of this study are available from the corresponding author TLD on request.

## References

1. Abecassis M, Bartlett ST, Collins AJ, et al. Kidney Transplantation as Primary Therapy for End-Stage Renal Disease: A National Kidney Foundation/Kidney Disease Outcomes Quality Initiative (NKF/KDOQI™) Conference. Clinical Journal of the American Society of Nephrology. 2008;3(2):471–480. doi:10.2215/cjn.05021107.

2. Rao PS, Ojo A. The Alphabet Soup of Kidney Transplantation: SCD, DCD, ECD—Fundamentals for the Practicing Nephrologist. Clinical Journal of the American Society of Nephrology. 2009;4(11):1827–1831. doi:10.2215/cjn.02270409.

3. Chopra B, Sureshkumar KK. Changing organ allocation policy for kidney transplantation in the United States. World Journal of Transplantation. 2015;5(2):38–43. doi:10.5500/wjt.v5.i2.38.

4. Organ Procurement and Transplantation Network. OPTN. Allocation Calculators. https://optn.transplant.hrsa.gov/resources/allocation-calculators/about-cpra/. Accessed May 2, 2020.

5. Wang CJ, Wetmore JB, Israni AK. Old versus new: Progress in reaching the goals of the new kidney allocation system. Human Immunology. 2017;78(1):9–15. doi:10.1016/j.humimm.2016.08.007.

6. Sanchez D, Dubay D, Prabhakar B, Taber DJ. Evolving Trends in Racial Disparities for Peri-Operative Outcomes with the New Kidney Allocation System (KAS) Implementation. Journal of Racial and Ethnic Health Disparities. 2018;5(6):1171–1179. doi:10.1007/s40615-018-0464-3.

7. Sethi S, Najjar R, Peng A, et al. Allocation of the Highest Quality Kidneys and Transplant Outcomes Under the New Kidney Allocation System. American Journal of Kidney Diseases. 2019;73(5):605–614. doi:10.1053/j.ajkd.2018.12.036.

8. Stewart DE, Wilk AR, Toll AE, et al. Measuring and monitoring equity in access to deceased donor kidney transplantation. American Journal of Transplantation. 2018;18(8):1924–1935. doi:10.1111/ajt.14922.

9. Barbas AS, Dib MJ, Rege AS, et al. The Volume-outcome Relationship in Deceased Donor Kidney Transplantation and Implications for Regionalization. Annals of Surgery. 2018;267(6):1169–1172. doi:10.1097/sla.0000000000002351.

10. Dahmen M, Becker F, Pavenstädt H, Suwelack B, Schütte-Nütgen K, Reuter S. Validation of the Kidney Donor Profile Index (KDPI) to assess a deceased donor’s kidneys’ outcome in a European cohort. Scientific Reports. 2019;9(1). doi:10.1038/s41598-019-47772-7.

11. Scientific Registry of Transplant Recipients: OPTN & SRTR 2011 Annual Data Report: Kidney. US Department of Health and Human Services, Health Resources and Services Administration. Available at: http://srtr.transplant.hrsa.gov/annual_reports/2011/flash/01_kidney/index.html. Accessed July 8, 2019

12. Kulkarni S, Ladin K, Haakinson D, Greene E, Li L, Deng Y. Association of Racial Disparities With Access to Kidney Transplant After the Implementation of the New Kidney Allocation System. JAMA Surgery. 2019;154(7):618–625. doi:10.1001/jamasurg.2019.0512.

13. Hickey MJ, Zheng Y, Valenzuela N, et al. New priorities: Analysis of the New Kidney Allocation System on UCLA patients transplanted from the deceased donor waitlist. Human Immunology. 2017;78(1):41–48. doi:10.1016/j.humimm.2016.10.020.

14. Vega WA, Rodriguez MA, Gruskin E. Health Disparities in the Latino Population. Epidemiologic Reviews. 2009;31(1):99–112. doi:10.1093/epirev/mxp008.

